# Quantifying body size estimation accuracy and body dissatisfaction in body dysmorphic disorder using a digital avatar

**DOI:** 10.1101/2024.02.21.24303163

**Authors:** Sameena Karsan, Joel P. Diaz-Fong, Ronald Ly, Gerhard Hellemann, Jamie D. Feusner

## Abstract

A core feature of body dysmorphic disorder (BDD) is body image disturbance. Many with BDD misperceive and are dissatisfied with the sizes and shapes of body parts, but detailed quantification and analysis of this has not yet been performed. To address this gap, we applied *Somatomap 3D,* a digital avatar tool, to quantify body image disturbances by assessing body size estimation (BSE) accuracy and body dissatisfaction. Sixty-one adults (31 with BDD, 30 healthy controls) created avatars to reflect their perceived current body and ideal body by altering 23 body part sizes and lengths using *Somatomap 3D.* Physical measurements of corresponding body parts were recorded for comparison. BSE accuracy (current minus actual) and body dissatisfaction (ideal minus current) were compared between groups and in relation to BDD symptom severity using generalized estimating equations. Individuals with BDD significantly over-and under-estimated certain body parts compared to healthy controls. Individuals with BDD overall desired significantly thinner body parts compared to healthy controls. Moreover, those with worse BSE accuracy had greater body dissatisfaction and poorer insight. In sum, this digital avatar tool revealed disturbances in body image in individuals with BDD that may have perceptual and cognitive/affective components.

## Introduction

Body dysmorphic disorder (BDD) is characterized by preoccupation with perceived defects of one’s appearance that are not noticeable or appear slight to others (Bjornsson et al., 2010; Phillips, 2004). This is associated with repetitive and avoidant behaviours and causes significant distress, functional impairment, and in some cases, suicidal ideation (Bjornsson et al., 2010; Eskander et al., 2020; Phillips & Menard, 2006a). Poor insight and delusionality of beliefs, where one is convinced that they have noticeable and prominent flaws with their appearance, are common in those with BDD (Phillips et al., 2006b; Phillips et al., 2010; Phillips et al., 2014). Approximately 2% of the general population is impacted by this psychiatric disorder (Fletcher, 2021; McGrath et al., 2023), with about 60-80% developing suicidal ideation and most having diminished quality of life (Bjornsson et al., 2010; Eskander et al., 2020; Phillips et al., 2006a; Phillips et al., 2008).

While the most common troublesome appearance features for those with BDD pertain to the face and head, appearance preoccupations involving the body – from the neck down – are also highly prevalent (Phillips, 2004; Phillips, 2005). Supporting this, 42% of total appearance concerns in a sample of individuals with BDD were of aspects of their body (from the neck down) (Phillips, 2005). Additionally, 65% of a BDD sample recruited for having primary face concerns reported having both face and body concerns (Moody et al., 2021). In another sample, 29% of participants with BDD had weight concerns (Kittler et al., 2007). Body concerns in those with BDD are frequently about the size of body parts being too large or too small. However, not all of these appearance concerns are related to body parts that are affected by weight gain or fat deposition (i.e., abdomen and thighs); these can also pertain to asymmetry, muscularity, excessive or not enough body hair, the size of genitals, body parts being too long or wide, etc. (Phillips, 2005).

The experience of those with BDD perceiving appearance features as defective when others do not notice or perceive them as slight imperfections may be due to distorted visual perception (Beilharz et al., 2017; Feusner et al., 2010; Jefferies et al., 2012; Toh et al., 2017a; Wong et al., 2022). Although, to date, research on visual perception in BDD has mainly focused on viewing and processing face stimuli. Further, in addition to visual sensory system disturbances, body size/shape misperceptions may have other mechanistic determinants, which could include disturbances in proprioception, somatosensory input, and/or multisensory integration (Kaplan et al., 2013), similar to what has been proposed in those with anorexia nervosa and bulimia nervosa, who also experience body image disturbance (Malighetti et al., 2022). Yet, little is still known about the underlying determinants of distorted body perception in those with BDD.

Current assessments of body image disturbance in individuals with BDD involve either clinician-administered scales or self-rated scales that gather information on one’s behaviours, cognitions, affect, and insight about their perceived physical appearance flaws. Although beneficial for detecting the presence of BDD symptoms and measuring their severity in general, these written and verbal language-based assessments fail to capture the perceptual experiences related to body image disturbance. Body image disturbance may arise from body dissatisfaction and/or body size distortion (inaccurate body size estimation; Hosseini & Padhy, 2023). Body size distortion is difficult to convey verbally or using rating scales and existing measures do not always allow for body part-specific descriptions of dissatisfaction. As such, current assessments may not comprehensively capture the manifestations of body image disturbance in BDD related to how an individual perceives themself.

*Somatomap 3D* is a digital avatar tool that was developed to quantitatively assess body image disturbance (Ralph-Nearman et al., 2019). With *Somatomap 3D* users can adjust the size and shape of individual body parts on a 3-dimensional avatar to a) indicate their best estimation of their current body part sizes, which can be used to determine their body size estimation (BSE) accuracy, and b) indicate the desired sizes and shapes of individual body parts, as a way of quantifying the degree of dissatisfaction. *Somatomap 3D* is sensitive to detecting abnormalities in BSE accuracy and body dissatisfaction for specific body parts in both nonclinical and clinical populations. Professional fashion models, using *Somatomap 3D*, had more accurate BSE accuracy than non-model controls (Ralph-Nearman et al., 2019). The application was also tested in individuals with anorexia nervosa, who were less accurate at estimating their body size compared to healthy controls and demonstrated greater body dissatisfaction for multiple body parts (Ralph-Nearman et al., 2021). Given the phenomenological overlaps between anorexia nervosa and BDD (Phillipou et al., 2019), including prominent body image disturbance, this tool may be suitable for measuring similar disturbances in BDD. Yet, no previous studies have investigated BSE accuracy or body dissatisfaction using *Somatomap 3D*, or other digital avatar tools, in those with BDD.

The present study had two primary goals: a) a proof of concept of using *Somatomap 3D* to detect BSE accuracy and to quantify body part dissatisfaction in a BDD population, and b) a comparison of BSE accuracy and body dissatisfaction between those with BDD and healthy controls. Given the phenomenological overlap related to perceptual distortions and body image disturbance in those with anorexia nervosa and those with BDD, we hypothesized that those with BDD would have lower BSE accuracy and greater body dissatisfaction compared to healthy controls. We also hypothesized that across participants, BSE accuracy would be associated with body dissatisfaction, such that more inaccurate estimations would be associated with greater dissatisfaction. Additionally, as more disturbed body perception could potentially lead to worse symptoms, we hypothesized that individuals with lower BSE accuracy would have worse preoccupations/repetitive behaviours and/or poorer insight. Finally, as body dissatisfaction could additionally be associated with worse BDD symptoms, we hypothesized that individuals with worse body dissatisfaction would also have worse preoccupations/repetitive behaviours and/or poorer insight.

## Material and methods

### Participants

This study was approved by the University of California, Los Angeles Institutional Review Board (17-000746, 2017) and the Centre for Addiction and Mental Health Research Ethics Board (075-2021, 2022). Prior to enrolment, participants provided informed consent. Unmedicated adults with BDD and healthy controls between the ages of 18-40 were recruited from the Greater Los Angeles Area (site 1) and Greater Toronto Area (site 2). Participants met with a clinician who performed a structured clinical interview. Participants included those with BDD and healthy controls (HCs) who participated in an ongoing study of visual processing. Individuals with BDD had primary concerns about their face and body and met DSM-5 criteria for BDD. Comorbid depressive, anxiety, or obsessive-compulsive and related disorders were allowed, which commonly co-occur in this population (Gunstad & Phillips, 2003), but those with other current comorbid psychiatric (including eating disorders) or substance use disorders were excluded. Those with lifetime bipolar disorder or psychotic disorders were also excluded. Healthy controls were excluded if they met criteria for any current DSM-5 disorders or lifetime bipolar disorder or psychotic disorders. Across all participants, exclusion criteria included suicidality, self-injurious behaviours, lifetime neurological disorder, current pregnancy, any medical illness that could affect cerebral metabolism, psychiatric medications in the prior eight weeks, or current treatment with cognitive-behavioural therapy. In addition, we measured visual acuity using a Snellen close vision eye chart to ensure that participants’ vision (corrected or uncorrected) was no worse than 20/30 in each eye.

### Diagnostic Evaluation and Symptom Assessment

Individuals who met the criteria for BDD on the BDD Diagnostic Module for DSM-5 (Phillips, 2017) and who scored ≥20 on the BDD version of the Yale–Brown Obsessive–Compulsive Scale (BDD-YBOCS) (Phillips et al., 1997) were eligible for inclusion in the BDD cohort. The BDD-YBOCS is a 12-item clinician administered assessment used to measure BDD severity. The Mini International Neuropsychiatric Inventory was administered to all participants to determine comorbid diagnoses (Sheehan et al., 1998). Participants who received a diagnosis of BDD during their structured clinical assessment completed the Brown Assessment of Beliefs Scale (BABS) (Eisen et al., 1998). The BABS is a 7-item clinician-administered assessment that measures insight about appearance beliefs. Higher scores indicate poorer insight (Eisen et al., 1998). All participants completed the 6-item self-rated Body Image States Scale (BISS) which measures one’s appraisal of their appearance at a certain moment (Cash et al., 2002). Scores range from one to nine with higher scores indicating positive body image.

### Somatomap 3D Procedures

Participants used *Somatomap 3D* (Figure 1) to engage with a three-dimensional avatar on a computer screen, as previously described (Ralph-Nearman et al., 2019; Ralph-Nearman et al., 2021). They were instructed to modify the avatar using sliders for 23 different body parts to create the sizes/shapes that best represented their perceived a) *current body*, and b) *ideal body*. For their *current body,* participants were instructed to “configure the body (from the neck down) to best match your current body shape”, and for their *ideal body* they were instructed to “configure the body (from the neck down) to best create the body size and shape that most closely matches with your ideal body.” The body parts included: neck girth and length, shoulder width, bust girth, chest girth, upper arm girth and length, lower arm girth and length, wrist girth, hand girth and length, torso length, waist size, stomach form, thigh girth and length, calf girth and length, ankle girth, and foot width and length. Participants were given 10 minutes to complete each portion of the *Somatomap 3D* tasks and were given a warning when they approached the 7-minute mark. The *Somatomap 3D* application was hosted on the secure Chorus app platform (https://www.joinchorus.com; Arevian et al., 2020).

**Figure 1.**
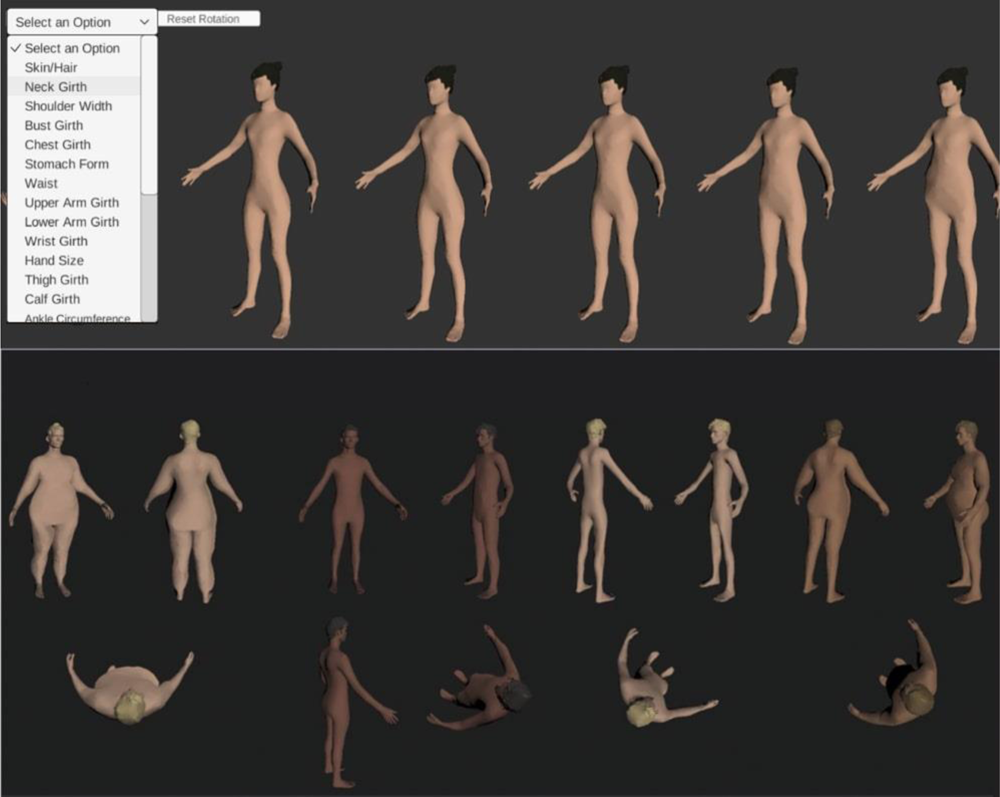
Example of *Somatomap 3D* interface. Participants could choose between a male and female avatar and then could alter their hair and skin color. The left drop-down menu has the list of 23 different body parts that could be altered. Avatars could be rotated along the x and y axis to view all angles of different body parts.

### Body Measurement Procedures

Research staff obtained physical measurements of the corresponding body parts after the *Somatomap 3D* procedures to minimize the possibility of bias during the tasks in the event participants inadvertently saw their measurements. Physical measurements were taken using a tape measure and were completed twice for the left and right side where applicable and then averaged. See https://dx.doi.org/10.17504/protocols.io.e6nvwdpr2lmk/v1 for a detailed description of how each body part was measured. Height and weight measurements were recorded using a stadiometer and a calibrated scale, respectively.

### Statistical analysis

Statistical analyses were performed with IBM SPSS Statistics (Version 27). Body measurements from *Somatomap 3D* were converted into centimetres from arbitrary units using previously described methods (Ralph-Nearman et al., 2019). A score for BSE accuracy was calculated for each body part by subtracting the physical body part measurement from one’s estimated body part size obtained from *Somatomap*. Body dissatisfaction was calculated by subtracting estimated measurements from ideal body part sizes. Before analyses, normality and homoscedasticity were assessed through visual inspection. Additionally, outliers were removed (0.8%) using Tukey’s method (Jones, 2019). An alpha level of .05 was used to determine statistical significance unless otherwise indicated. We used study site as a covariate for all analyses.

We compared overall and body part-specific BSE accuracy and dissatisfaction between groups for the 23 body parts using generalized estimating equations (GEEs), to account for potentially correlated values across body parts for BSE and/or dissatisfaction. The dependent variables for the models were BSE accuracy and dissatisfaction while the independent variables were group, body part, and a group X body part interaction effect variable. We also examined if there was a relationship between BSE accuracy and body dissatisfaction using a GEE. This model was similar to the model previously described, but BSE was added as an additional main effect, as well as group X BSE and group X body part X BSE interaction effects. Covariates of non-interest included site, height, weight, and body mass index (BMI). We did not control for gender since, from previous analyses, men and women did not show different patterns once height, weight, and BMI were controlled for (Guo et al., 2023).

To assess the association between body image disturbances and BDD symptom severity, for the BDD group, a) BSE accuracy was entered into separate GEE models with the BDD-YBOCS and BABS as dependent variables and b) body dissatisfaction was entered into separate GEE models with the BDD-YBOCS and BABS as dependent variables. To test each of the separate hypotheses, we used a corrected alpha significance level of .025 to account for the two BDD symptom severity measure dependent variables.

We also performed an exploratory analysis across the whole sample (BDD and HC) to assess the associations between BSE accuracy and/or body dissatisfaction with body image appraisal as measured by the BISS, using similar GEE models.

## Results

### Participants

A total of 61 adult unmedicated participants (31 with BDD and 30 HCs) completed the study (Table 1). Sixty participants completed ratings of BSE while 40 participants completed ratings of body dissatisfaction. Thirty-nine participants completed both ratings. Forty-two participants were from the greater Los Angeles area and 19 were from the Greater Toronto Area. The majority of participants (82%) were women. The mean age for individuals with BDD was 23.40 ± 5.63 and HCs was 22.87 ± 6.20. BDD participants had significantly lower (worse) appraisals of their body image (M = 3.28, SD = 1.17) compared to HCs (M = 6.07, SD = 1.19) as measured by the BISS [*t*(55) = 8.90, *p* <.001]. Table 1 contains full demographic and psychometric characteristics.

**Table 1:**
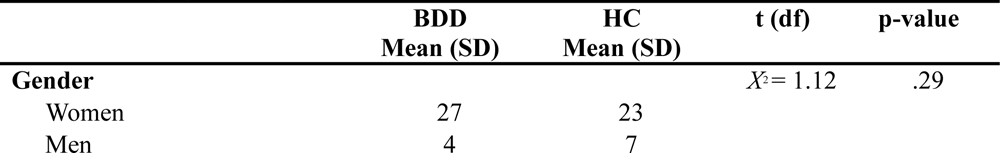
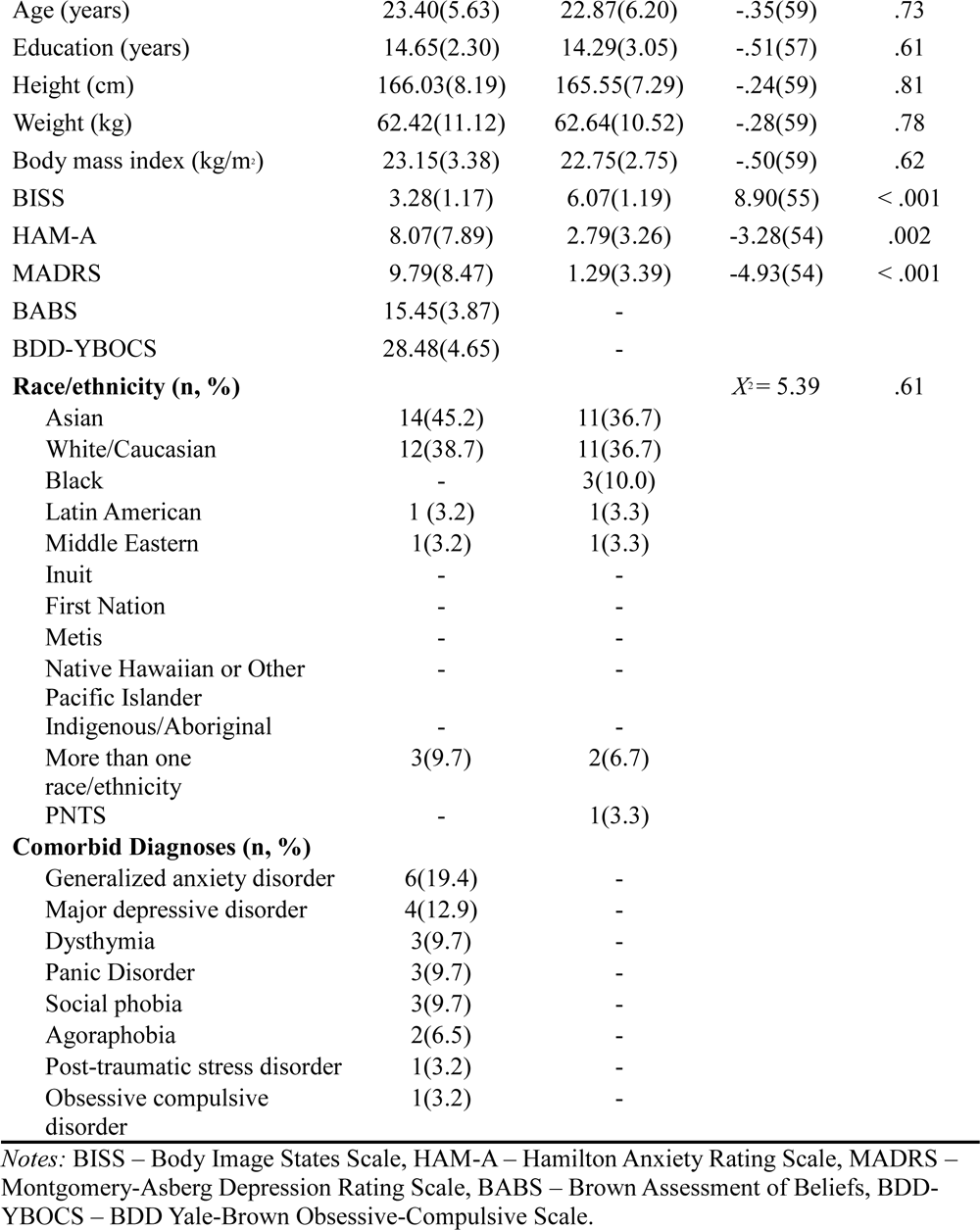
Demographic and psychometric characteristics.

|  | **BDD**<br>**Mean (SD)** | **HC**<br>**Mean (SD)** | **t (df)** | **p-value** |
| --- | --- | --- | --- | --- |
| **Gender** | | | $\chi^2 = 1.12$ | .29 |
| Women | 27 | 23 |  |  |
| Men | 4 | 7 |  |  |
| Age (years) | 23.40(5.63) | 22.87(6.20) | -.35(59) | .73 |
| Education (years) | 14.65(2.30) | 14.29(3.05) | -.51(57) | .61 |
| Height (cm) | 166.03(8.19) | 165.55(7.29) | -.24(59) | .81 |
| Weight (kg) | 62.42(11.12) | 62.64(10.52) | -.28(59) | .78 |
| Body mass index (kg/m<sup>2</sup>) | 23.15(3.38) | 22.75(2.75) | -.50(59) | .62 |
| BISS | 3.28(1.17) | 6.07(1.19) | 8.90(55) | < .001 |
| HAM-A | 8.07(7.89) | 2.79(3.26) | -3.28(54) | .002 |
| MADRS | 9.79(8.47) | 1.29(3.39) | -4.93(54) | < .001 |
| BABS | 15.45(3.87) | - |  |  |
| BDD-YBOCS | 28.48(4.65) | - |  |  |
| **Race/ethnicity (n, %)** | | | $\chi^2 = 5.39$ | .61 |
| Asian | 14(45.2) | 11(36.7) |  |  |
| White/Caucasian | 12(38.7) | 11(36.7) |  |  |
| Black | - | 3(10.0) |  |  |
| Latin American | 1 (3.2) | 1(3.3) |  |  |
| Middle Eastern | 1(3.2) | 1(3.3) |  |  |
| Inuit | - | - |  |  |
| First Nation | - | - |  |  |
| Metis | - | - |  |  |
| Native Hawaiian or Other<br>Pacific Islander | - | - |  |  |
| Indigenous/Aboriginal | - | - |  |  |
| More than one<br>race/ethnicity | 3(9.7) | 2(6.7) |  |  |
| PNTS | - | 1(3.3) |  |  |
| **Comorbid Diagnoses (n, %)** |  |  |  |  |
| Generalized anxiety disorder | 6(19.4) | - |  |  |
| Major depressive disorder | 4(12.9) | - |  |  |
| Dysthymia | 3(9.7) | - |  |  |
| Panic Disorder | 3(9.7) | - |  |  |
| Social phobia | 3(9.7) | - |  |  |
| Agoraphobia | 2(6.5) | - |  |  |
| Post-traumatic stress disorder | 1(3.2) | - |  |  |
| Obsessive compulsive<br>disorder | 1(3.2) | - |  |  |

*Notes:* BISS – Body Image States Scale, HAM-A – Hamilton Anxiety Rating Scale, MADRS – Montgomery-Asberg Depression Rating Scale, BABS – Brown Assessment of Beliefs, BDD-YBOCS – BDD Yale-Brown Obsessive-Compulsive Scale.

### BSE Accuracy

The main effect of group for BSE accuracy was not significant (Wald *X*^2^ [1, *N =* 60] = .35, *p* = .55); however, there was a significant main effect of body part (Wald *X*^2^ [22, *N =* 60] = 545.47, *p <* .001); both groups underestimated the sizes of most body parts. There was also a significant interaction effect of group X body part (Wald *X*^2^ [22, *N =* 60] = 41.65, *p =* .007; Figure 2), that is, BSE accuracy differed between groups depending on the body part being estimated. Specifically, the groups differed in their ability to estimate their lower arm girth size (*p* = .04); those with BDD overestimated (M = -3.35, SE = .83) compared to controls (M = -5.34, SE = .47). Additionally, estimation of bust girth and foot width showed trends toward significance between groups (*p*s < .10); those with BDD underestimated both their bust girth (M = -3.75, SE = 1.59) compared to HC (M = .26, SE = 1.74) and their foot width (M = -.72, SE = .59) compared to HCs (M = .44, SE = .37).

**Figure 2.**
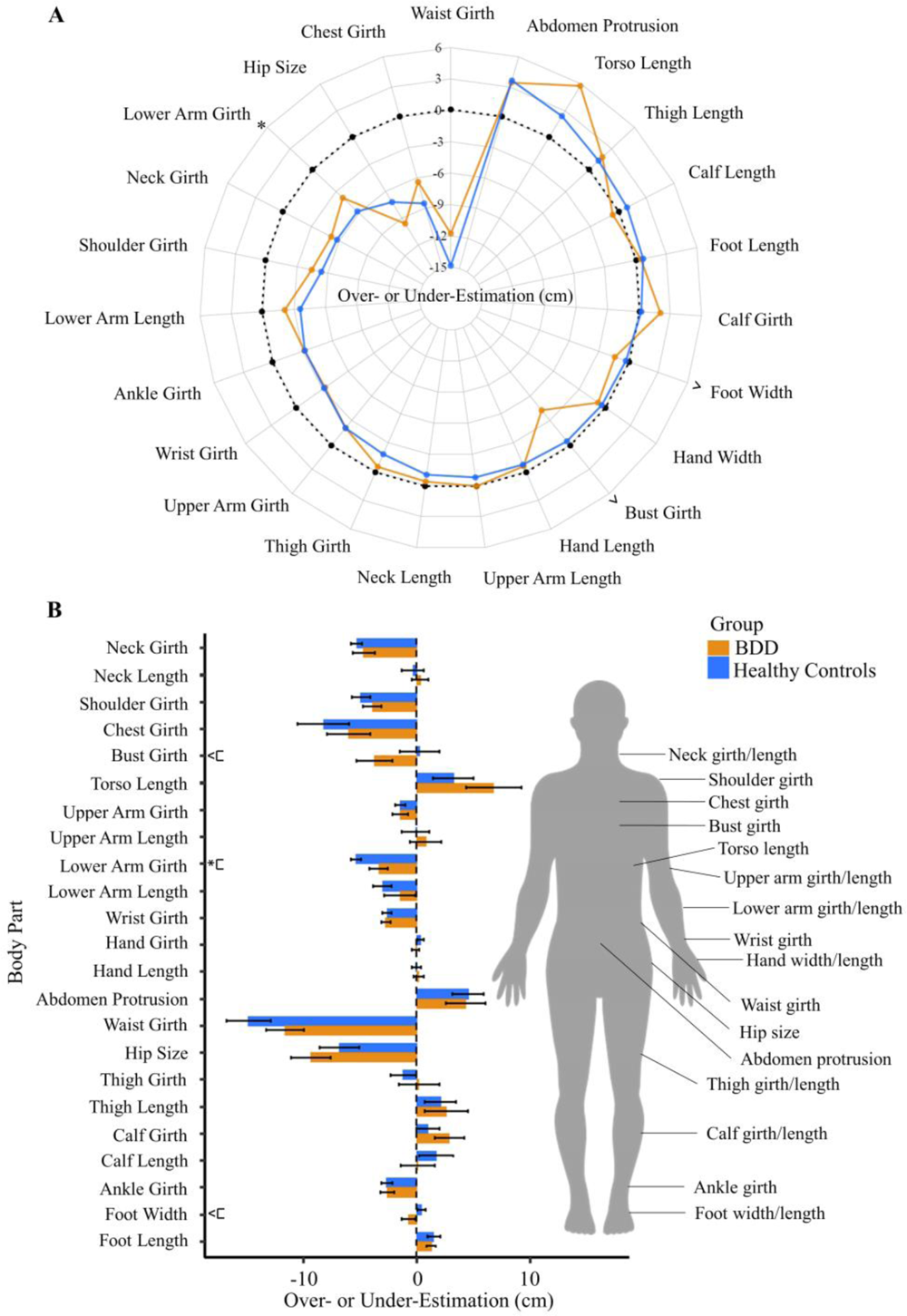
Body size estimation (BSE) inaccuracy (cm) in body dysmorphic disorder (BDD) and healthy control participants. (A) Radar chart has body parts arranged clockwise in the order of overestimation to underestimation in healthy controls and (B) bar chart has body parts organized from top of the body to the body and has standard error bars. The manikin on right indicates the body parts that are manipulated during the *Somatomap 3D* task. Positive values indicate overestimation of body part size, negative values indicate underestimation, and zero indicates perfect accuracy. * *p* < .05, ^ *p <* .100

### Body Dissatisfaction

There was a significant difference between groups for body dissatisfaction (Wald *X*^2^ [1, *N =* 40] = 8.93, *p =* .003); those with BDD, on average, desired smaller body parts. There was also a significant main effect of body part (Wald *X*^2^ [22, *N =* 40] = 282.38, *p* < .001) and interaction of group X body part (Wald *X*^2^ [22, *N =* 40] = 48.64, *p* < .001; Figure 3). Pertaining to the latter, those with BDD desired smaller calf (*p* = .04), chest (*p* < .001), lower arm (*p* = .007), upper arm (*p* = .04), and thigh girths (*p* = .007); and lower abdomen protrusion (*p* = .005), compared to controls. Dissatisfaction with upper arm length and shoulder width showed a trend toward significance between groups (*p*s ≤ .100).

**Figure 3.**
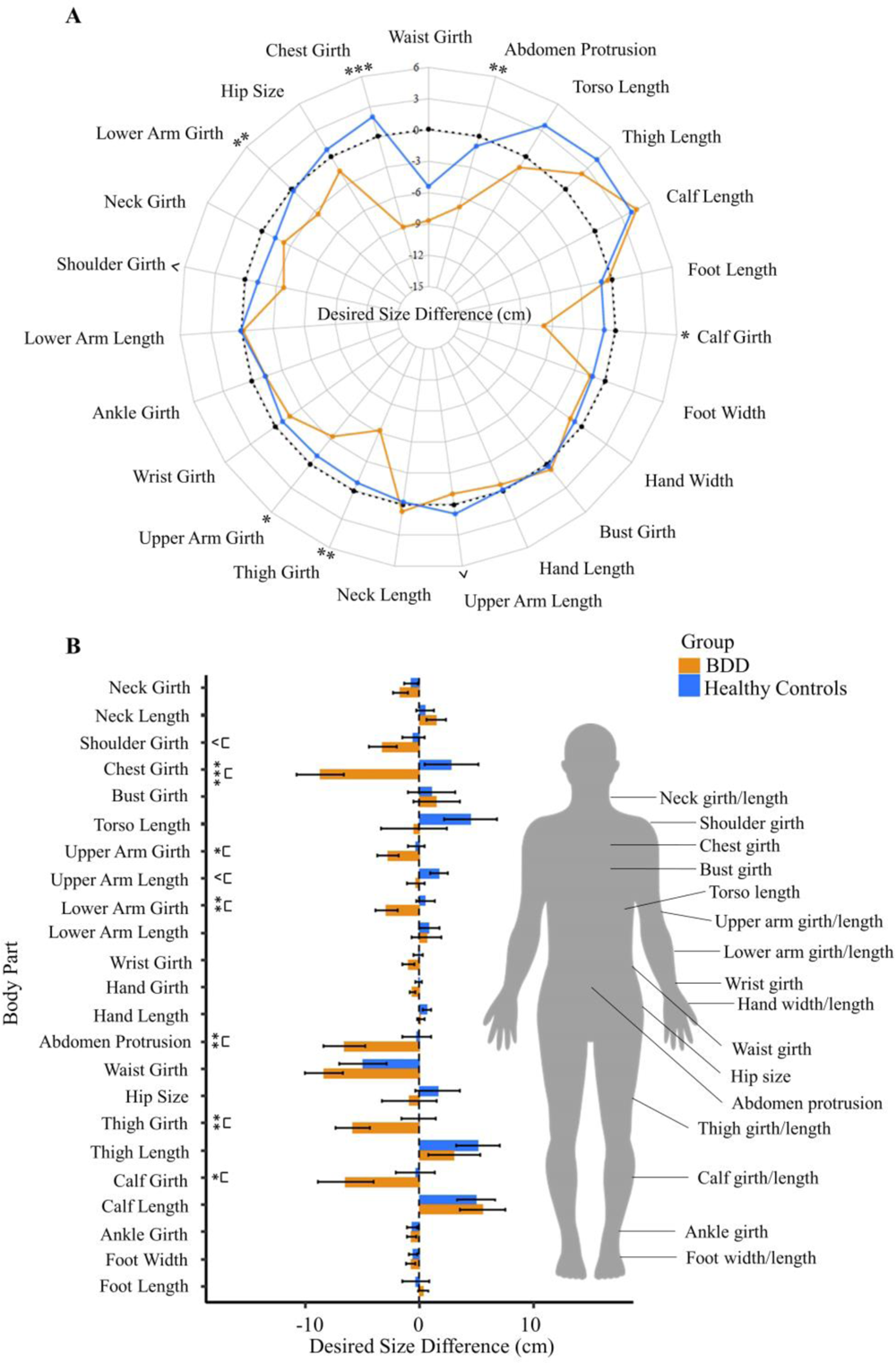
Desired body size difference (cm) (“dissatisfaction”) between body dysmorphic disorder (BDD) and healthy control participants. Desired size for each body part was calculated by the difference between the ideal avatar and the estimated current body size avatar. (A) Radar chart and (B) Bar chart has standard error bars. The manikin on right indicates the body parts that are manipulated during the *Somatomap 3D* task. Positive values indicate desiring a larger body part size, negative values indicate desiring a smaller body part. Values closer to zero indicate greater satisfaction. ^ *p <* .10; * *p <* .05, ** *p <* .01, *** *p <* .001

### Relationships between BSE Accuracy and Body Dissatisfaction

BSE accuracy and body dissatisfaction were significantly associated (Wald *X*^2^ [1, *N =* 39] = 78.44, *p <* .001). The main effect of group remained significant in this updated model (Wald *X*^2^ [1, *N =* 39] = 9.73, *p =* .002). Significant interactions of group X body part (Wald *X*^2^ [22, *N =* 39] = 57.71, *p <* .001) and group X body part X BSE (Wald *X*^2^ [38, *N =* 39] = 2.30E5, *p <* .001) were observed, but the group X BSE interaction was not significant (Wald *X*^2^ [1, *N =* 39] = 1.28, *p =* .26). A post-hoc analysis revealed that the association between BSE accuracy and body dissatisfaction differed between groups for bust girth (*p* = .03), calf girth (*p* = .05), foot length (*p* = .03), hand length (*p* = .001), and lower arm girth (*p* = .001). Bust, calf, and lower arm girths followed the same pattern in which greater inaccuracy at estimating girth was related to greater dissatisfaction for those with BDD but not controls. The pattern for foot and hand length, however, was significant for healthy controls but not BDD (Figure 4).

**Figure 4.**
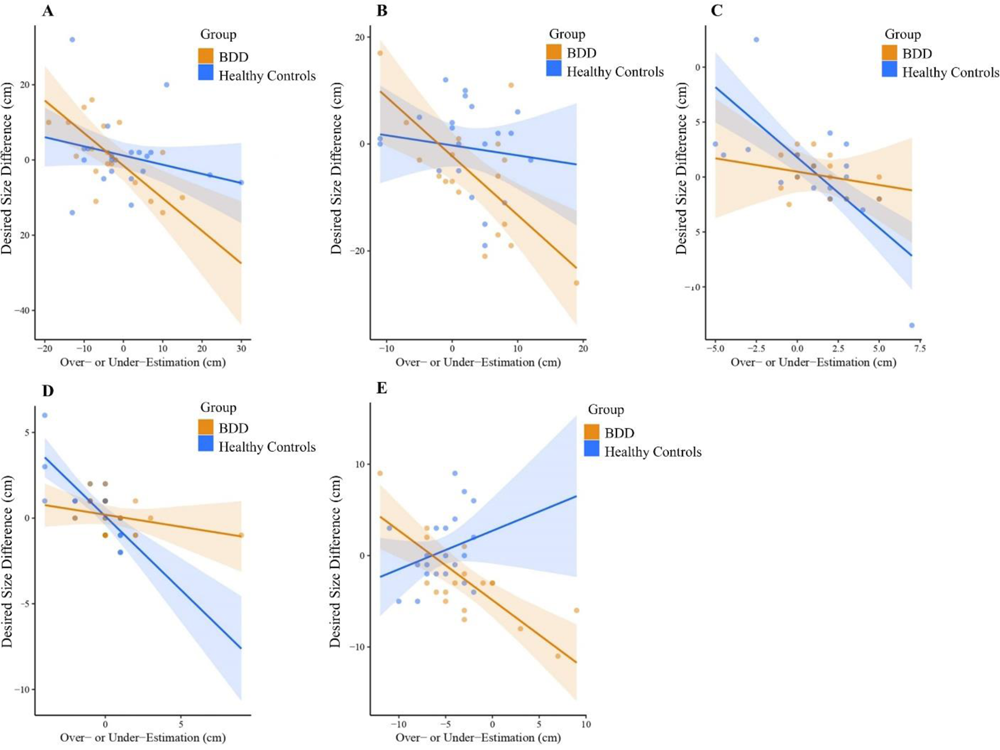
Group differences for the relationship between body dissatisfaction and body size estimation (BSE) inaccuracy. Out of the 23 body parts, significant group X BSE X body part interactions were observed for (A) bust girth, (B) calf girth, (C) foot length, (D) hand length, and (E) lower arm girth. Individuals with BDD showed a significant relation between body dissatisfaction and BSE accuracy for bust, calf, and lower arm girths, such that greater inaccuracy was related to greater dissatisfaction. Healthy controls, but not individuals with BDD, showed the same significant association for foot and hand lengths.

### BDD-YBOCS and BABS

In individuals with BDD, BSE accuracy was associated with BABS scores (Wald *X*^2^ [1, *N =* 30] = 6.17, *p* = .013); overestimation of body size was related to worse insight (β = .05). BDD-YBOCS was not significantly associated with BSE accuracy or body dissatisfaction (Table 2).

**Table 2:** Generalized estimating equation (GEE) regression coefficients for BSE accuracy and body dissatisfaction predicting BDD symptom severity in the BDD group using the BDD-YBOCS and BABS.

| Variable | BDD-YBOCS |  |  |  | BABS |  |  |  |
| --- | --- | --- | --- | --- | --- | --- | --- | --- |
| | $\beta$ | SE | 95% CI | $p$ | $\beta$ | SE | 95% CI | $p$ |
| BSE Accuracy | .01 | .03 | -.05, .07 | .74 | .05 | .02 | .01, .09 | **.01** |
| Body Dissatisfaction | -.09 | .05 | -.18, .01 | .07 | -.02 | .04 | -.11, .06 | .61 |

Notes: BDD-YBOCS – BDD Yale-Brown Obsessive-Compulsive Scale, BABS – Brown Assessment of Beliefs.

### Exploratory and Post-hoc Analyses

BISS scores were not significantly associated with BSE accuracy or body dissatisfaction (Table S5).

We conducted exploratory post-hoc correlation analyses using absolute residual values from the group GEE models and BDD symptom severity measures to assess whether individuals who exhibited greater variation from the group were associated with worse pathology. No findings were significant, suggesting no association within the current sample (Table S5).

Sensitivity analyses were completed without participants who identified as men (n = 4 BDD; n = 7 HCs) to see if the same results held. All results remained the same; however, the group difference for body dissatisfaction showed a trend towards significance [Wald *X*^2^ (1, *N =* 34) = 3.60, *p* = .058] and the same was observed for BSE accuracy predicting BABS scores [Wald *X*^2^ (1, *N =* 27) = 4.59, *p* = .032]. Results for all exploratory analyses can be viewed in the supplemental section (Tables S6 to S10).

## Discussion

This study quantified disturbances in body image in individuals with BDD using *Somatomap 3D* and examined their relationships to clinical symptoms. While both groups tended to underestimate the size of most body parts, those with BDD showed several significant body part-specific over-or under-estimations compared with HCs. Body dissatisfaction showed a more consistent pattern, in which those with BDD desired most body parts to be smaller compared with HCs. Importantly, there were strong relationships between BSE accuracy and body dissatisfaction, which were more pronounced for certain body parts and were differential by group. Further, those with BDD with worse BSE accuracy had worse insight.

### Those with BDD show abnormal BSE

*Somatomap 3D* detected differences in those with BDD compared with HCs in the ability to accurately estimate the size/shape of body parts. This effect was limited to specific body parts, similar to what was previously demonstrated in those with anorexia nervosa (Ralph-Nearman et al., 2021). Unlike the findings in anorexia nervosa (Ralph-Nearman et al., 2021) in which these body parts were significantly misestimated to be larger (mostly corresponding to body areas that are more sensitive to fat deposition such as lower abdominal protrusion, hip size, and thigh and calf girths), individuals with BDD showed either over-or under-estimations depending on the body part (Figure 2). While both disorders share a core diagnostic feature of body image disturbance, comparison studies highlight different patterns of areas of concern among individuals with anorexia nervosa and BDD, potentially related to variations in BSE accuracy. Specifically, those with anorexia nervosa have prominent concerns about being overweight and having a larger body width/girth in areas that are sensitive to fat deposition, whereas individuals with BDD have a wider range of concerns including body asymmetry, skin, and hair (Rosen & Ramirez, 1998; Toh et al., 2020). Moreover, an exclusion criterion of the present study was comorbid eating disorders. Speculatively, individuals with BDD with comorbid eating disorders might have similar patterns of overestimating their body size, as what is typically observed in individuals with primary anorexia nervosa.

Abnormal visual processing in BDD may contribute to BSE inaccuracies. Individuals with BDD show selective visual attention biases (Greenberg et al., 2014; Grocholewski et al., 2012; Kollei et al., 2017; Toh et al., 2017b), greater sensitivity to details that may not be apparent to others (Feusner et al., 2010a; Toh et al., 2017a), disturbances in functional neural systems responsible for global visual perception for viewing own-and others’ faces as well as houses and bodies (Feusner et al., 2007; Feusner et al., 2010b, Feusner et al., 2011, Li et al., 2015a, Li et al., 2015b Moody et al., 2021,Wong et al., 2021 Wong et al., 2022), and disturbances in white-and gray matter structure in visual systems (Arienzo et al., 2013; Feusner et al., 2013, Feusner et al., 2023). These factors can enter a feedback loop with compulsive behaviours such as checking the mirror frequently and fixating on body parts, or else completely avoiding mirrors and abstaining from looking at their body (Castle et al., 2021), all of which may contribute to further attentional biases. The distorted visual percept resulting from this, and associated visual memories, could thus impact their representation of their internal body size and lead to greater inaccuracies in BSE. In addition to the impact of abnormal visual perception, abnormalities in other sensory modalities (such as somatosensation, proprioception, equilibrioception, interoception, and/or multisensory integration) may contribute to inaccurate BSE and body dissatisfaction. A mismatch between how one would expect their body to feel in a situation and how it actually feels could create a sense of “body mistrust,” possibly impacting one’s ability to estimate their body size accurately (Jenkinson & Rossell, 2023), although this has not yet been directly tested in individuals with BDD. To shed light on this, future research could simultaneously measure visual attention and visual and other sensory modality perception when completing BSE tasks.

BSE inaccuracy in those with BDD showed a significant relationship with poor insight (BABS scores), as hypothesized, but, unexpectedly, not with preoccupations/repetitive behaviours (BDD-YBOCS scores). Also contrary to predictions, dissatisfaction was not significantly associated with either the BDD-YBOCS or BABS. Further, there were no significant associations across the whole sample between BSE or dissatisfaction with appearance appraisals (BISS scores). Thus, the key emergent clinical association finding is that those with BDD with worse BSE accuracy have poorer insight. This may reflect previously observed associations between worse insight and disturbances in visual processing-related brain function and structure (Feusner et al., 2013; Li et al., 2015). Disturbances in visual processing may contribute (along with, hypothetically, disturbances in other sensory modalities such as proprioception, somatosensation, or multisensory integration) to misperceived body part sizes/shapes, and hence poorer accuracy in BSE. The worse these disturbances are, the harder they may be for the individual to refute since most tend, by default, to trust their sensory experiences. This has important clinical implications, as those with poorer insight have a lower likelihood of seeking mental health treatment (Phillips et al., 2005).

The control group, on average, underestimated the size of most body parts, which is consistent with previous studies examining body size estimation patterns in healthy controls using different assessment methods (Drumond Andrade et al., 2009; Gruszka et al., 2022; Kapka-Skrzypczak et al., 2012; Ralph-Nearman et al., 2019). However, if healthy individuals typically underestimate specific body parts, it would logically follow that *more accurate* estimations in these areas could indicate pathology since it would represent overestimation relative to healthy norms. Future research should examine BSE patterns in large, population-based samples using tools that allow alteration of individual body parts to establish normative body size representation and how it may vary with features including demographics, geographical location of residence, body morphometrics, etc. This could be used in comparison with clinical samples to discern deviance from normative body size representations, suggesting the presence of BSE pathology.

### Those with BDD show a high degree of body dissatisfaction across many body parts

A more consistent pattern of group differences in the current study was observed for body dissatisfaction. Those with BDD exhibited greater dissatisfaction overall with their body, mostly desiring smaller body parts. Some, but not all, of the body parts that the BDD group was dissatisfied with – lower arm girth – were ones in which they had abnormal BSE accuracy. Chest, thigh and calf girths, and lower abdomen protrusion, had the largest magnitude of differences in dissatisfaction between groups. Patterns of body dissatisfaction may be more impacted by sociocultural zeitgeists than body size estimation. Media representation of an ideal body type (Bennett et al., 2020; Marques et al., 2022), cultural-based differential influences (Santhira Shagar et al., 2021), and peer influences or family dynamics (Helfert & Warschburger, 2011; Michael et al., 2014) have been documented to contribute to the development of body dissatisfaction.

### Impairment in BSE accuracy is associated with body dissatisfaction across the sample

BSE accuracy and body dissatisfaction showed significant relationships across BDD and healthy controls, demonstrating this association regardless of the presence of BDD. However, the significant three-way interaction between group, body part, and BSE accuracy when predicting body dissatisfaction underscores the complex interplay between these variables. Specifically, within the BDD group a significant relationship between BSE accuracy and body dissatisfaction was observed for girths only (bust, calf, and lower arm) whereas in healthy controls this relationship was significant for lengths (foot and hand). One’s mental representation of their body and the way they consistently perceive themself, mainly their beliefs about their body, may lead to dissatisfaction (Heider et al., 2018). Conversely, body dissatisfaction, which is characterized by negative attitudes toward one’s appearance, may predict distortions in how one perceives their body and estimates of their body size/shape (Tremblay & Limbos, 2009). The directionality of these relationships remains to be determined; however, both variables could reinforce one another creating a harmful cycle that can foster body image disturbances and lead to the progression of disorders such as BDD and anorexia nervosa (Alexi et al., 2019; Möllmann et al., 2024). As such, future research should focus on disentangling this relationship through longitudinal intervention studies to discern which factor precedes the other and how this can be leveraged for treatments.

### Clinical implications

There are several clinical implications of these findings. Individuals with BDD often do not receive appropriate treatment or are misdiagnosed (Schulte et al., 2020). Many seek cosmetic treatments, as they believe that the root of their problems is a physical aspect of their appearance, yet are often still dissatisfied after the procedures (Lai et al., 2010; Veale, 2000). This includes high rates of cosmetic procedures to alter body parts such as breast reconstruction, lipectomy, and liposuction (Lee et al., 2023). However, cosmetic surgery is contraindicated for this population as it is typically associated with poor outcomes and does not reduce BDD symptom severity (Bowyer et al., 2016). The current findings provide quantitative evidence of body part size misestimation in those with BDD, including a trend for underestimation of bust girth, which is a common procedure. This may explain why cosmetic surgery largely does not improve BDD symptoms in this population since their inability to accurately assess the size of certain body parts (linked to their dissatisfaction), might persist after surgery. This remains to be directly tested, however.

Further, while cognitive behavioural therapy (CBT) and pharmacological treatments can be effective treatments for BDD, there are several barriers associated with them. Particularly, many with BDD believe that mental health treatment would be ineffective since it does not address what they think is their core problem as they are strongly convinced their flaws are real and visible (Marques et al., 2011), i.e, they have poor insight. In a clinical setting, a visual tool such as *Somatomap 3D* might assist clinicians and other mental health professionals in improving insight by demonstrating to patients the discrepancies between their body part size estimations and their actual, measured body part sizes. In addition, such a visual tool could help clinicians and clinical researchers engage with patients to catalogue and track body parts they are dissatisfied with over the course of treatment. Future studies employing longitudinal designs could test the degree *Somatomap* is sensitive to detecting changes in BDD severity naturalistically over time (e.g. in at-risk populations in childhood through adolescence) and as the result of treatment interventions. In sum, *Somatomap 3D* might help clinicians better understand a patient’s internal body image, track progress over time, and tailor treatment strategies to specific areas of concern that cause distress.

### Limitations

There are several limitations that should be considered. Importantly, participants in this study took part in a larger study of visual processing of faces, whose inclusion criteria included having primary face concerns. Thus, while all included in this analysis had both face and body concerns, the sample may not be as representative as one that also included those with *only* body concerns. It is possible that those with primary body concerns could manifest more extreme dissatisfaction and possibly worse BSE, although this remains to be tested. Another limitation is that the majority of the participants in this sample were women (82%). Yet, BDD also impacts a large proportion of men (∼40%; Phillips & Castle, 2001). To ensure the generalizability of results, future studies should aim to have a balanced representation of genders. Another consideration is that specific body part appearance appraisals are quite varied and idiosyncratic across individuals with BDD, and for some body parts there is no dissatisfaction. Similarly, BSE accuracy for some body parts (as evidenced in this study) is similar to that in controls. Thus, quantification of *average* degrees of dissatisfaction, and BSE accuracy, for each body part at the group level is susceptible to random sampling biases. A much larger study would be necessary to better delineate, for example, clusters of individuals with patterns of sets of body parts with more or less dissatisfaction or BSE accuracy. The current analyses, nevertheless, provide meaningful general patterns of differences between those with BDD and healthy controls.

## Conclusions

In sum, these results demonstrate disturbances in BSE accuracy and body dissatisfaction in individuals with BDD using a digital avatar tool. Those with BDD may exhibit abnormalities in size estimations of certain body parts, which may be due to distorted perception and disturbances in their internal body image. Further, they show greater dissatisfaction than controls across many body parts, desiring most parts to be smaller. *Somatomap 3D* shows promise as a visually based tool for assessing body part dissatisfaction and for assessing abnormalities in BSE related to internal body image in individuals with BDD.

## CRediT Authorship Contribution Statement

**Sameena Karsan:** Formal analysis, Writing – original draft, Writing – review & editing, Visualization. **Joel P. Diaz-Fong:** Investigation, Data curation, Writing – review & editing. **Ronald Ly:** Investigation, Data curation, Writing – review & editing. **Gerhard Hellemann:** Methodology, Writing – review & editing. **Jamie D. Feusner:** Conceptualization, Methodology, Writing – review & editing, Supervision, Project administration.

## Data Availability

Data is available from the corresponding author upon reasonable request.

## Acknowledgements

The Software used in this research was created by Jamie D. Feusner, Armen C, Arevian, and Nanthia Suthana (UCLA); and Sahib S. Khalsa and Christina Ralph-Nearman (Laureate Institute for Brain Research). © 2023 UCLA. Originally published in JMIR Mental Health (http://mental.jmir.org), 29.10.2019. Ralph-Nearman, C., Arevian, A. C., Puhl, M., Kumar, R., Villaroman, D., Suthana, N., Feusner, J. D., & Khalsa, S. S. (2019). A Novel Mobile Tool (Somatomap) to Assess Body Image Perception Pilot Tested With Fashion Models and Nonmodels: Cross-Sectional Study. JMIR mental health, 6(10), e14115. https://doi.org/10.2196/14115 Commercial entities: please contact for licensing opportunities.

## Notes

### Competing Interest Statement

The authors have declared no competing interest.

### Clinical Trial

NCT04373629

### Funding Statement

This work was funded by the National Institute of Mental Health (NIMH) grant numbers: R21MH110865 (Feusner) & R01MH121520 (Feusner).

### Author Declarations

The University of California, Los Angeles Institutional Review Board (17-000746, 2017) and the Centre for Addiction and Mental Health Research Ethics Board (075-2021, 2022) gave ethical approval for this work.

