## Supplemental Tables for "Quantifying body size estimation accuracy and body dissatisfaction in body dysmorphic disorder using a digital avatar"

Table S1: Parameter estimates for body size estimation (BSE) inaccuracy generalized estimating equation (GEE) model

| *Predictors* | Estimates | Standard Error | 95% Confidence Interval | | *p*-value |
| --- | --- | --- | --- | --- | --- |
|  |  |  | Lower Bound | Upper Bound |  |
| (Intercept) | -36.51 | 15.17 | -66.25 | -6.78 | **0.02** |
| Group [control] | .11 | 0.58 | -1.02 | 1.25 | 0.84 |
| Body Part [Ankle Girth] | .14 | 0.51 | -0.87 | 1.15 | 0.79 |
| Body Part [Bust Girth] | -1.00 | 1.62 | -4.18 | 2.17 | 0.54 |
| Body Part [Calf Girth] | 5.62 | 1.27 | 3.13 | 8.12 | **< .001** |
| Body Part [Calf Length] | 2.85 | 1.50 | -0.10 | 5.79 | 0.06 |
| Body Part [Chest Girth] | -3.26 | 1.85 | -6.89 | 0.37 | 0.08 |
| Body Part [Foot Length] | 4.02 | 0.62 | 2.80 | 5.24 | **< .001** |
| Body Part [Foot Width] | 2.02 | 0.58 | 0.89 | 3.15 | **< .001** |
| Body Part [Hand Length] | 2.88 | 0.53 | 1.83 | 3.93 | **< .001** |
| Body Part [Hand Width] | 2.64 | 0.45 | 1.75 | 3.53 | **< .001** |
| Body Part [Hip Size] | -6.61 | 1.76 | -10.06 | -3.17 | **< .001** |
| Body Part [Lower Arm Girth] | -0.62 | 0.70 | -1.99 | 0.76 | 0.38 |
| Body Part [Lower Arm Length] | 1.26 | 1.33 | -1.35 | 3.86 | 0.34 |
| Body Part [Neck Girth] | -1.93 | 0.88 | -3.65 | -0.21 | **0.03** |
| Body Part [Neck Length] | 3.08 | 0.76 | 1.58 | 4.57 | **< .001** |
| Body Part [Shoulder Width] | -1.19 | 0.99 | -3.12 | 0.74 | 0.23 |
| Body Part [Abdomen Protrusion] | 7.06 | 1.71 | 3.70 | 10.41 | **< .001** |
| Body Part [Thigh Girth] | 2.95 | 1.67 | -0.33 | 6.22 | 0.08 |
| Body Part [Thigh Length] | 5.35 | 1.77 | 1.89 | 8.82 | **0.002** |
| Body Part [Torso Height] | 9.52 | 2.37 | 4.88 | 14.17 | **< .001** |
| Body Part [Upper Arm Girth] | 1.27 | 0.68 | -0.06 | 2.61 | 0.06 |
| Body Part [Upper Arm Length] | 3.55 | 1.33 | 0.93 | 6.17 | **0.01** |
| Body Part [Waist Size] | -8.88 | 1.64 | -12.10 | -5.66 | **< .001** |
| Body Part [Wrist Girth] | - | - | - | - | - |
| Group [control]*Body Part [Ankle Girth] | -0.15 | 0.79 | -1.69 | 1.40 | 0.85 |
| Group [control]*Body Part [Bust Girth] | 3.89 | 2.34 | -0.69 | 8.47 | 0.10 |
| Group [control]*Body Part [Calf Girth] | -2.04 | 1.58 | -5.13 | 1.06 | 0.20 |
| Group [control]*Body Part [Calf Length] | 1.49 | 2.10 | -2.62 | 5.61 | 0.48 |
| Group [control]*Body Part [Chest Girth] | -2.35 | 2.86 | -7.96 | 3.26 | 0.41 |
| Group [control]*Body Part [Foot Length] | 0.12 | 0.96 | -1.77 | 2.00 | 0.90 |
| Group [control]*Body Part [Foot Width] | 1.05 | 0.71 | -0.34 | 2.44 | 0.14 |
| Group [control]*Body Part [Hand Length] | -0.29 | 0.71 | -1.68 | 1.11 | 0.69 |
| Group [control]*Body Part [Hand Width] | 0.32 | 0.59 | -0.84 | 1.49 | 0.59 |
| Group [control]*Body Part [Hip Size] | 2.41 | 2.42 | -2.34 | 7.16 | 0.32 |
| Group [control]*Body Part [Lower Arm Girth] | -2.10 | 0.84 | -3.74 | -0.46 | **0.01** |
| Group [control]*Body Part [Lower Arm Length] | -1.66 | 1.59 | -4.79 | 1.46 | 0.30 |
| Group [control]*Body Part [Neck Girth] | -0.76 | 1.08 | -2.88 | 1.37 | 0.49 |
| Group [control]*Body Part [Neck Length] | -0.80 | 1.36 | -3.48 | 1.87 | 0.56 |
| Group [control]*Body Part [Shoulder Width] | -1.09 | 1.33 | -3.71 | 1.52 | 0.41 |
| Group [control]*Body Part [Abdomen Protrusion] | 0.10 | 2.27 | -4.34 | 4.54 | 0.97 |
| Group [control]*Body Part [Thigh Girth] | -1.49 | 2.01 | -5.43 | 2.44 | 0.46 |
| Group [control]*Body Part [Thigh Length] | -0.64 | 2.34 | -5.22 | 3.94 | 0.78 |
| Group [control]*Body Part [Torso Height] | -3.65 | 2.98 | -9.49 | 2.19 | 0.22 |
| Group [control]*Body Part [Upper Arm Girth] | -0.10 | 0.89 | -1.84 | 1.65 | 0.92 |
| Group [control]*Body Part [Upper Arm Length] | -1.01 | 1.92 | -4.78 | 2.76 | 0.60 |
| Group [control]*Body Part [Waist Size] | -3.33 | 2.51 | -8.25 | 1.58 | 0.18 |
| Group [control]*Body Part [Wrist Girth] | - | - | - | - | - |
| Site [CAMH] | -0.23 | 0.70 | -1.60 | 1.14 | 0.74 |
| Height (m) | 0.23 | 0.10 | 0.03 | 0.43 | 0.03 |
| Weight (kg) | -0.27 | 0.09 | -0.46 | -0.09 | 0.004 |
| BMI (kg/m^2^) | 0.56 | 0.20 | 0.16 | 0.96 | 0.01 |

*Notes:* Wrist girth set to zero since parameter is redundant. BMI = body mass index. Site, height, weight, and BMI were covariates in the model.

Table S2: Parameter estimates for body dissatisfaction generalized estimating equation (GEE) model

| *Predictors* | Estimates | Standard Error | 95% Confidence Interval | | | *p*-value |
| --- | --- | --- | --- | --- | --- | --- |
|  |  |  | Lower Bound | | Upper Bound |  |
| (Intercept) | 73.07 | 60.70 | -45.91 | 192.05 | | 0.23 |
| Group [control] | 0.90 | 0.65 | -0.37 | 2.18 | | 0.17 |
| Body Part [Ankle Girth] | 0.32 | 0.59 | -0.83 | 1.47 | | 0.59 |
| Body Part [Bust Girth] | 2.47 | 2.19 | -1.83 | 6.78 | | 0.26 |
| Body Part [Calf Girth] | -5.47 | 2.43 | -10.25 | -0.70 | | **0.03** |
| Body Part [Calf Length] | 6.53 | 1.93 | 2.74 | 10.32 | | **0.001** |
| Body Part [Chest Girth] | -7.68 | 1.88 | -11.36 | -4.01 | | **<.001** |
| Body Part [Foot Length] | 1.34 | 0.63 | 0.10 | 2.58 | | **0.03** |
| Body Part [Foot Width] | 0.24 | 0.53 | -0.81 | 1.28 | | 0.66 |
| Body Part [Hand Length] | 1.11 | 0.57 | -0.01 | 2.23 | | 0.05 |
| Body Part [Hand Width] | 0.40 | 0.49 | -0.56 | 1.35 | | 0.42 |
| Body Part [Hip Size] | 0.11 | 2.55 | -4.90 | 5.11 | | 0.97 |
| Body Part [Lower Arm Girth] | -1.90 | 0.92 | -3.69 | -0.10 | | **0.04** |
| Body Part [Lower Arm Length] | 1.63 | 1.37 | -1.05 | 4.31 | | 0.23 |
| Body Part [Neck Girth] | -0.68 | 0.72 | -2.10 | 0.73 | | 0.34 |
| Body Part [Neck Length] | 2.47 | 1.00 | 0.51 | 4.44 | | **0.01** |
| Body Part [Shoulder Width] | -2.21 | 1.41 | -4.97 | 0.54 | | 0.12 |
| Body Part [Abdomen Protrusion] | -5.58 | 1.97 | -9.44 | -1.72 | | **0.01** |
| Body Part [Thigh Girth] | -4.84 | 1.59 | -7.96 | -1.73 | | **0.002** |
| Body Part [Thigh Length] | 4.05 | 2.16 | -0.18 | 8.29 | | 0.06 |
| Body Part [Torso Height] | 0.53 | 2.96 | -5.28 | 6.34 | | 0.86 |
| Body Part [Upper Arm Girth] | -1.79 | 0.82 | -3.41 | -0.17 | | **0.03** |
| Body Part [Upper Arm Length] | 0.68 | 0.81 | -0.89 | 2.26 | | 0.40 |
| Body Part [Waist Size] | -7.37 | 1.69 | -10.69 | -4.05 | | **<.001** |
| Body Part [Wrist Girth] | - | - | - | - | | - |
| Group [control]*Body Part [Ankle Girth] | -0.87 | 0.85 | -2.54 | 0.79 | | 0.31 |
| Group [control]*Body Part [Bust Girth] | -1.28 | 2.96 | -7.08 | 4.51 | | 0.66 |
| Group [control]*Body Part [Calf Girth] | 5.24 | 3.05 | -0.73 | 11.21 | | 0.09 |
| Group [control]*Body Part [Calf Length] | -1.48 | 2.51 | -6.40 | 3.44 | | 0.56 |
| Group [control]*Body Part [Chest Girth] | 10.59 | 3.07 | 4.58 | 16.60 | | **0.001** |
| Group [control]*Body Part [Foot Length] | -1.56 | 1.40 | -4.30 | 1.19 | | 0.27 |
| Group [control]*Body Part [Foot Width] | -0.69 | 0.62 | -1.90 | 0.52 | | 0.26 |
| Group [control]*Body Part [Hand Length] | -0.34 | 0.67 | -1.66 | 0.97 | | 0.61 |
| Group [control]*Body Part [Hand Width] | -0.37 | 0.66 | -1.66 | 0.92 | | 0.57 |
| Group [control]*Body Part [Hip Size] | 1.61 | 3.18 | -4.62 | 7.84 | | 0.61 |
| Group [control]*Body Part [Lower Arm Girth] | 2.51 | 1.23 | 0.10 | 4.93 | | **0.04** |
| Group [control]*Body Part [Lower Arm Length] | -0.68 | 1.67 | -3.96 | 2.60 | | 0.69 |
| Group [control]*Body Part [Neck Girth] | 0.07 | 0.98 | -1.85 | 1.98 | | 0.95 |
| Group [control]*Body Part [Neck Length] | -1.86 | 1.33 | -4.46 | 0.75 | | 0.16 |
| Group [control]*Body Part [Shoulder Width] | 1.78 | 1.76 | -1.66 | 5.22 | | 0.31 |
| Group [control]*Body Part [Abdomen Protrusion] | 5.44 | 2.39 | 0.76 | 10.11 | | **0.02** |
| Group [control]*Body Part [Thigh Girth] | 4.84 | 2.22 | 0.49 | 9.19 | | **0.03** |
| Group [control]*Body Part [Thigh Length] | 1.19 | 2.97 | -4.63 | 7.00 | | 0.69 |
| Group [control]*Body Part [Torso Height] | 4.05 | 3.74 | -3.28 | 11.37 | | 0.28 |
| Group [control]*Body Part [Upper Arm Girth] | 1.60 | 1.17 | -0.69 | 3.89 | | 0.17 |
| Group [control]*Body Part [Upper Arm Length] | 1.13 | 1.15 | -1.13 | 3.38 | | 0.33 |
| Group [control]*Body Part [Waist Size] | 2.51 | 2.61 | -2.61 | 7.63 | | 0.34 |
| Group [control]*Body Part [Wrist Girth] | - | - | - | - | | - |
| Height | -0.42 | 0.37 | -1.14 | 0.31 | | 0.26 |
| Weight | 0.53 | 0.46 | -0.38 | 1.43 | | 0.25 |
| BMI | -1.68 | 1.27 | -4.17 | 0.82 | | 0.19 |

*Notes:* Wrist girth set to zero since parameter is redundant. Height, weight, and BMI were covariates in the model.

Table S3: Parameter estimates for body dissatisfaction generalized estimating equation (GEE) with body size estimation (BSE) in the model

| *Predictors* | Estimates | Standard Error | 95% Confidence Interval | | | *p*-value |
| --- | --- | --- | --- | --- | --- | --- |
|  |  |  | Lower Bound | Upper Bound | |  |
| (Intercept) | 6.56 | 59.09 | -109.26 | | 122.38 | 0.91 |
| Group [control] | 1.50 | 1.03 | -0.52 | | 3.51 | 0.15 |
| BSE | -0.58 | 0.22 | -1.01 | | -0.15 | **0.01** |
| Group [control]*BSE | 0.25 | 0.26 | -0.26 | | 0.76 | 0.33 |
| Body Part [Ankle Girth] | 1.35 | 0.67 | 0.04 | | 2.66 | **0.04** |
| Body Part [Bust Girth] | 1.49 | 1.54 | -1.53 | | 4.51 | 0.33 |
| Body Part [Calf Girth] | -0.37 | 2.00 | -4.28 | | 3.54 | 0.85 |
| Body Part [Calf Length] | 8.20 | 1.79 | 4.69 | | 11.71 | **<.001** |
| Body Part [Chest Girth] | -8.24 | 2.46 | -13.06 | | -3.42 | **<.001** |
| Body Part [Foot Length] | 3.79 | 1.09 | 1.66 | | 5.92 | **<.001** |
| Body Part [Foot Width] | 1.85 | 0.89 | 0.11 | | 3.59 | **0.04** |
| Body Part [Hand Length] | 2.95 | 0.80 | 1.39 | | 4.52 | **<.001** |
| Body Part [Hand Width] | 1.98 | 0.77 | 0.47 | | 3.49 | **0.01** |
| Body Part [Hip Size] | -5.61 | 3.03 | -11.56 | | 0.33 | 0.06 |
| Body Part [Lower Arm Girth] | -2.18 | 0.53 | -3.22 | | -1.14 | **<.001** |
| Body Part [Lower Arm Length] | 1.45 | 1.25 | -0.99 | | 3.90 | 0.24 |
| Body Part [Neck Girth] | 0.15 | 0.75 | -1.32 | | 1.62 | 0.84 |
| Body Part [Neck Length] | 4.50 | 1.04 | 2.46 | | 6.54 | **<.001** |
| Body Part [Shoulder Width] | -3.62 | 1.63 | -6.82 | | -0.41 | **0.03** |
| Body Part [Abdomen Protrusion] | -1.99 | 1.89 | -5.69 | | 1.71 | 0.29 |
| Body Part [Thigh Girth] | -3.93 | 1.14 | -6.16 | | -1.70 | **<.001** |
| Body Part [Thigh Length] | 6.80 | 2.82 | 1.28 | | 12.33 | **0.02** |
| Body Part [Torso Height] | 6.84 | 2.29 | 2.34 | | 11.33 | **<.001** |
| Body Part [Upper Arm Girth] | -2.17 | 1.22 | -4.56 | | 0.21 | **0.07** |
| Body Part [Upper Arm Length] | 3.18 | 1.03 | 1.16 | | 5.19 | **<.001** |
| Body Part [Waist Size] | -9.49 | 2.14 | -13.68 | | -5.29 | **<.001** |
| Body Part [Wrist Girth] | - | - | - | | - | - |
| Group [control]*Body Part [Ankle Girth] | -2.44 | 1.18 | -4.76 | | -0.12 | **0.04** |
| Group [control]*Body Part [Bust Girth] | 0.73 | 2.43 | -4.03 | | 5.49 | 0.76 |
| Group [control]*Body Part [Calf Girth] | 1.61 | 2.63 | -3.55 | | 6.76 | 0.54 |
| Group [control]*Body Part [Calf Length] | -2.46 | 2.39 | -7.15 | | 2.24 | 0.31 |
| Group [control]*Body Part [Chest Girth] | 10.61 | 3.27 | 4.20 | | 17.01 | **<.001** |
| Group [control]*Body Part [Foot Length] | -1.43 | 1.37 | -4.10 | | 1.25 | 0.30 |
| Group [control]*Body Part [Foot Width] | -1.09 | 1.11 | -3.27 | | 1.10 | 0.33 |
| Group [control]*Body Part [Hand Length] | -1.72 | 1.01 | -3.70 | | 0.25 | 0.09 |
| Group [control]*Body Part [Hand Width] | -1.12 | 1.03 | -3.15 | | 0.91 | 0.28 |
| Group [control]*Body Part [Hip Size] | 4.46 | 3.78 | -2.94 | | 11.86 | 0.24 |
| Group [control]*Body Part [Lower Arm Girth] | 5.81 | 2.03 | 1.82 | | 9.80 | **<.001** |
| Group [control]*Body Part [Lower Arm Length] | -1.96 | 2.01 | -5.90 | | 1.98 | 0.33 |
| Group [control]*Body Part [Neck Girth] | -1.59 | 1.97 | -5.44 | | 2.26 | 0.42 |
| Group [control]*Body Part [Neck Length] | -3.23 | 1.46 | -6.09 | | -0.37 | **0.03** |
| Group [control]*Body Part [Shoulder Width] | 2.48 | 2.29 | -2.00 | | 6.96 | 0.28 |
| Group [control]*Body Part [Abdomen Protrusion] | 3.19 | 2.33 | -1.37 | | 7.75 | 0.17 |
| Group [control]*Body Part [Thigh Girth] | 3.25 | 2.04 | -0.75 | | 7.25 | 0.11 |
| Group [control]*Body Part [Thigh Length] | 0.02 | 3.50 | -6.84 | | 6.87 | .99 |
| Group [control]*Body Part [Torso Height] | 1.10 | 2.81 | -4.41 | | 6.60 | 0.70 |
| Group [control]*Body Part [Upper Arm Girth] | 2.55 | 1.68 | -0.74 | | 5.83 | 0.13 |
| Group [control]*Body Part [Upper Arm Length] | -0.55 | 1.41 | -3.33 | | 2.22 | 0.70 |
| Group [control]*Body Part [Waist Size] | 0.73 | 3.25 | -5.65 | | 7.11 | 0.82 |
| Group [control]*Body Part [Wrist Girth] | - | - | - | | - | - |
| Group [control]*Body Part [Ankle Girth]*BSE | -0.19 | 0.24 | -0.66 | | 0.28 | 0.43 |
| Group [control]*Body Part [Bust Girth]*BSE | 0.15 | 0.27 | -0.38 | | 0.67 | 0.59 |
| Group [control]*Body Part [Calf Girth]*BSE | 0.04 | 0.23 | -0.42 | | 0.49 | 0.87 |
| Group [control]*Body Part [Calf Length]*BSE | -0.07 | 0.19 | -0.45 | | 0.30 | 0.70 |
| Group [control]*Body Part [Chest Girth]*BSE | 0.12 | 0.21 | -0.28 | | 0.52 | 0.56 |
| Group [control]*Body Part [Foot Length]*BSE | -0.64 | 0.45 | -1.51 | | 0.24 | 0.16 |
| Group [control]*Body Part [Foot Width]*BSE | -0.08 | 0.17 | -0.42 | | 0.26 | 0.66 |
| Group [control]*Body Part [Hand Length]*BSE | -0.31 | 0.25 | -0.79 | | 0.18 | 0.22 |
| Group [control]*Body Part [Hand Width]*BSE | -0.44 | 0.30 | -1.02 | | 0.14 | 0.14 |
| Group [control]*Body Part [Hip Size]*BSE | -0.04 | 0.18 | -0.40 | | 0.32 | 0.83 |
| Group [control]*Body Part [Lower Arm Girth]*BSE | 0.73 | 0.33 | 0.08 | | 1.38 | **0.03** |
| Group [control]*Body Part [Lower Arm Length]*BSE | -0.18 | 0.25 | -0.67 | | 0.31 | 0.47 |
| Group [control]*Body Part [Neck Girth]*BSE | 0.07 | 0.35 | -0.63 | | 0.76 | 0.85 |
| Group [control]*Body Part [Neck Length]*BSE | 0.10 | 0.18 | -0.25 | | 0.46 | 0.57 |
| Group [control]*Body Part [Shoulder Width]*BSE | -0.09 | 0.22 | -0.53 | | 0.35 | 0.69 |
| Group [control]*Body Part [Abdomen Protrusion]*BSE | 0.20 | 0.21 | -0.21 | | 0.60 | 0.35 |
| Group [control]*Body Part [Thigh Girth]*BSE | -0.27 | 0.28 | -0.82 | | 0.28 | 0.34 |
| Group [control]*Body Part [Thigh Length]*BSE | 0.09 | 0.22 | -0.35 | | 0.53 | 0.69 |
| Group [control]*Body Part [Torso Height]*BSE | -0.36 | 0.20 | -0.74 | | 0.02 | 0.07 |
| Group [control]*Body Part [Upper Arm Girth]*BSE | 0.19 | 0.34 | -0.47 | | 0.85 | 0.57 |
| Group [control]*Body Part [Upper Arm Length]*BSE | 0.23 | 0.13 | -0.02 | | 0.48 | 0.08 |
| Group [control]*Body Part [Waist Size]*BSE | 0.08 | 0.17 | -0.25 | | 0.41 | 0.63 |
| Group [control]*Body Part [Wrist Girth]*BSE | - | - | - | | - | - |
| Height (m) | -0.01 | 0.36 | -0.71 | | 0.70 | 0.98 |
| Weight (kg) | 0.07 | 0.45 | -0.82 | | 0.96 | 0.88 |
| BMI (kg/m^2^) | -0.52 | 1.25 | -2.97 | | 1.92 | 0.67 |

*Notes:* Wrist girth set to zero since parameter is redundant. Height, weight, and BMI were covariates in the model.

Table S4: GEE regression coefficients for BSE accuracy and body dissatisfaction predicting BDD symptom severity using the Body Image States Scale (BISS) across the full group

|  | **BISS** | | | |
| --- | --- | --- | --- | --- |
| **Variable** | β | SE | 95% CI | *p* |
| *Model 1* |  |  |  |  |
| BSE Accuracy | -.01 | .01 | -.03, .01 | .26 |
| Group | 2.79 | .31 | 2.19, 3.40 | **< .001** |
| Group*BSE Accuracy | .01 | .01 | -.04, .76 | .58 |
| *Model 2* |  |  |  |  |
| Body Dissatisfaction | .01 | .01 | -.02, .03 | .50 |
| Group | 2.99 | .42 | 2.18, 3.81 | **< .001** |
| Group*Body Dissatisfaction | .01 | .02 | -.03, .04 | .64 |

Table S5: Spearman correlations between residuals from GEE group model and BDD symptom severity

|  | BDD-YBOCS | BABS |
| --- | --- | --- |
| BSE Accuracy Residual | .25  (*p* = .19) | .19  (*p* = .32) |
| Body Dissatisfaction Residual | -.10  (*p* = .67) | .28  (*p* = .24) |

Table S6: Women only parameter estimates for body size estimation (BSE) inaccuracy generalized estimating equation (GEE) model

| *Predictors* | Estimates | Standard Error | 95% Confidence Interval | | *p*-value |
| --- | --- | --- | --- | --- | --- |
|  |  |  | Lower Bound | Upper Bound |  |
| (Intercept) |  |  |  |  |  |
| Group [control] | -51.34 | 15.12 | -80.97 | -21.72 | **0.001** |
| Body Part [Ankle Girth] | 0.49 | 0.53 | -0.55 | 1.52 | 0.35 |
| Body Part [Bust Girth] | -1.82 | 1.72 | -5.19 | 1.55 | 0.29 |
| Body Part [Calf Girth] | 5.34 | 1.32 | 2.75 | 7.93 | **< 0.001** |
| Body Part [Calf Length] | 3.29 | 1.60 | 0.16 | 6.43 | **0.04** |
| Body Part [Chest Girth] | -4.99 | 1.72 | -8.36 | -1.63 | **0.004** |
| Body Part [Foot Length] | 4.21 | 0.65 | 2.95 | 5.48 | **< 0.001** |
| Body Part [Foot Width] | 1.89 | 0.63 | 0.65 | 3.13 | **0.003** |
| Body Part [Hand Length] | 3.09 | 0.57 | 1.97 | 4.21 | **< 0.001** |
| Body Part [Hand Width] | 2.62 | 0.50 | 1.64 | 3.60 | **< 0.001** |
| Body Part [Hip Size] | -5.65 | 1.77 | -9.13 | -2.18 | **0.001** |
| Body Part [Lower Arm Girth] | -0.70 | 0.75 | -2.18 | 0.78 | 0.35 |
| Body Part [Lower Arm Length] | 0.67 | 1.09 | -1.47 | 2.82 | 0.54 |
| Body Part [Neck Girth] | -1.44 | 0.91 | -3.23 | 0.35 | 0.12 |
| Body Part [Neck Length] | 3.57 | 0.76 | 2.08 | 5.05 | **< 0.001** |
| Body Part [Shoulder Width] | -0.83 | 1.05 | -2.89 | 1.23 | 0.43 |
| Body Part [Abdomen Protrusion] | 8.01 | 1.78 | 4.53 | 11.49 | **< 0.001** |
| Body Part [Thigh Girth] | 2.09 | 1.73 | -1.30 | 5.48 | 0.23 |
| Body Part [Thigh Length] | 7.10 | 1.52 | 4.12 | 10.08 | **< 0.001** |
| Body Part [Torso Height] | 11.05 | 2.37 | 6.40 | 15.69 | **< 0.001** |
| Body Part [Upper Arm Girth] | 0.92 | 0.72 | -0.50 | 2.33 | 0.20 |
| Body Part [Upper Arm Length] | 5.06 | 1.14 | 2.83 | 7.28 | **< 0.001** |
| Body Part [Waist Size] | -7.98 | 1.73 | -11.38 | -4.58 | **< 0.001** |
| Body Part [Wrist Girth] | 0a | . | . | . | . |
| Group [control]*Body Part [Ankle Girth] | -0.50 | 0.75 | -1.96 | 0.97 | 0.51 |
| Group [control]*Body Part [Bust Girth] | 3.26 | 2.15 | -0.95 | 7.47 | 0.13 |
| Group [control]*Body Part [Calf Girth] | -1.61 | 1.66 | -4.87 | 1.66 | 0.33 |
| Group [control]*Body Part [Calf Length] | 1.84 | 2.26 | -2.59 | 6.27 | 0.42 |
| Group [control]*Body Part [Chest Girth] | -2.40 | 2.81 | -7.91 | 3.11 | 0.39 |
| Group [control]*Body Part [Foot Length] | 0.84 | 0.83 | -0.78 | 2.45 | 0.31 |
| Group [control]*Body Part [Foot Width] | 1.22 | 0.80 | -0.35 | 2.78 | 0.13 |
| Group [control]*Body Part [Hand Length] | 0.28 | 0.72 | -1.13 | 1.69 | 0.70 |
| Group [control]*Body Part [Hand Width] | 0.59 | 0.66 | -0.71 | 1.88 | 0.37 |
| Group [control]*Body Part [Hip Size] | 2.86 | 2.53 | -2.10 | 7.81 | 0.26 |
| Group [control]*Body Part [Lower Arm Girth] | -1.82 | 0.93 | -3.64 | 0.01 | 0.05 |
| Group [control]*Body Part [Lower Arm Length] | -0.56 | 1.50 | -3.50 | 2.38 | 0.71 |
| Group [control]*Body Part [Neck Girth] | 1.17 | 1.59 | -1.95 | 4.29 | 0.46 |
| Group [control]*Body Part [Neck Length] | 0.02 | 1.44 | -2.79 | 2.84 | 0.99 |
| Group [control]*Body Part [Shoulder Width] | -1.81 | 1.44 | -4.64 | 1.02 | 0.21 |
| Group [control]*Body Part [Abdomen Protrusion] | 2.44 | 2.03 | -1.55 | 6.42 | 0.23 |
| Group [control]*Body Part [Thigh Girth] | 0.31 | 2.08 | -3.77 | 4.40 | 0.88 |
| Group [control]*Body Part [Thigh Length] | -0.54 | 2.12 | -4.70 | 3.62 | 0.80 |
| Group [control]*Body Part [Torso Height] | -4.87 | 2.91 | -10.58 | 0.84 | 0.10 |
| Group [control]*Body Part [Upper Arm Girth] | 0.00 | 1.02 | -2.00 | 2.00 | 0.99 |
| Group [control]*Body Part [Upper Arm Length] | 0.33 | 1.56 | -2.74 | 3.39 | 0.84 |
| Group [control]*Body Part [Waist Size] | -4.82 | 2.67 | -10.04 | 0.41 | 0.07 |
| Group [control]*Body Part [Wrist Girth] | 0.49 | 0.53 | -0.55 | 1.52 | 0.35 |
| Site [CAMH] | -0.30 | 0.66 | -1.59 | 0.98 | 0.65 |
| Height (m) | 0.34 | 0.10 | 0.13 | 0.54 | **0.001** |
| Weight (kg) | -0.34 | 0.08 | -0.48 | -0.19 | **< 0.001** |
| BMI (kg/m^2^) | 0.62 | 0.15 | 0.33 | 0.92 | **< 0.001** |

*Notes:* Wrist girth set to zero since parameter is redundant. BMI = body mass index. Site, height, weight, and BMI were covariates in the model.

Table S7: Women only parameter estimates for body dissatisfaction generalized estimating equation (GEE) model

| *Predictors* | Estimates | Standard Error | 95% Confidence Interval | | | *p*-value |
| --- | --- | --- | --- | --- | --- | --- |
|  |  |  | Lower Bound | | Upper Bound |  |
| (Intercept) | 88.77 | 61.76 | -32.27 | 209.82 | | 0.15 |
| Group [control] | 0.69 | 0.72 | -0.71 | 2.10 | | 0.33 |
| Body Part [Ankle Girth] | 0.32 | 0.59 | -0.83 | 1.46 | | 0.59 |
| Body Part [Bust Girth] | 2.47 | 2.19 | -1.83 | 6.78 | | 0.26 |
| Body Part [Calf Girth] | -5.47 | 2.44 | -10.25 | -0.70 | | **0.03** |
| Body Part [Calf Length] | 6.53 | 1.93 | 2.74 | 10.32 | | **0.001** |
| Body Part [Chest Girth] | -7.68 | 1.88 | -11.36 | -4.01 | | **< 0.001** |
| Body Part [Foot Length] | 1.34 | 0.63 | 0.10 | 2.58 | | **0.03** |
| Body Part [Foot Width] | 0.24 | 0.53 | -0.81 | 1.28 | | 0.66 |
| Body Part [Hand Length] | 1.11 | 0.57 | -0.01 | 2.22 | | 0.05 |
| Body Part [Hand Width] | 0.39 | 0.49 | -0.56 | 1.35 | | 0.42 |
| Body Part [Hip Size] | 0.11 | 2.55 | -4.90 | 5.11 | | 0.97 |
| Body Part [Lower Arm Girth] | -1.89 | 0.92 | -3.69 | -0.10 | | **0.04** |
| Body Part [Lower Arm Length] | 1.63 | 1.37 | -1.05 | 4.31 | | 0.23 |
| Body Part [Neck Girth] | -0.68 | 0.72 | -2.10 | 0.73 | | 0.34 |
| Body Part [Neck Length] | 2.47 | 1.00 | 0.51 | 4.44 | | **0.01** |
| Body Part [Shoulder Width] | -2.21 | 1.41 | -4.97 | 0.54 | | 0.12 |
| Body Part [Abdomen Protrusion] | -5.58 | 1.97 | -9.44 | -1.72 | | **0.01** |
| Body Part [Thigh Girth] | -4.84 | 1.59 | -7.95 | -1.73 | | **0.002** |
| Body Part [Thigh Length] | 4.05 | 2.16 | -0.18 | 8.29 | | **0.06** |
| Body Part [Torso Height] | 0.53 | 2.96 | -5.28 | 6.34 | | 0.86 |
| Body Part [Upper Arm Girth] | -1.79 | 0.82 | -3.40 | -0.17 | | **0.03** |
| Body Part [Upper Arm Length] | 0.68 | 0.81 | -0.89 | 2.26 | | 0.40 |
| Body Part [Waist Size] | -7.37 | 1.69 | -10.69 | -4.05 | | **< 0.001** |
| Body Part [Wrist Girth] | - | - | - | - | | - |
| Group [control]*Body Part [Ankle Girth] | -0.92 | 0.89 | -2.65 | 0.82 | | 0.30 |
| Group [control]*Body Part [Bust Girth] | -3.41 | 2.67 | -8.64 | 1.83 | | 0.20 |
| Group [control]*Body Part [Calf Girth] | 3.81 | 3.15 | -2.36 | 9.97 | | 0.23 |
| Group [control]*Body Part [Calf Length] | -1.93 | 2.75 | -7.31 | 3.46 | | 0.48 |
| Group [control]*Body Part [Chest Girth] | 7.75 | 2.64 | 2.57 | 12.93 | | **0.003** |
| Group [control]*Body Part [Foot Length] | -1.11 | 0.90 | -2.88 | 0.66 | | 0.22 |
| Group [control]*Body Part [Foot Width] | -0.60 | 0.61 | -1.80 | 0.59 | | 0.32 |
| Group [control]*Body Part [Hand Length] | -0.57 | 0.68 | -1.91 | 0.77 | | 0.40 |
| Group [control]*Body Part [Hand Width] | -0.43 | 0.68 | -1.76 | 0.90 | | 0.53 |
| Group [control]*Body Part [Hip Size] | 1.36 | 3.26 | -5.04 | 7.76 | | 0.68 |
| Group [control]*Body Part [Lower Arm Girth] | 0.76 | 1.16 | -1.52 | 3.04 | | 0.51 |
| Group [control]*Body Part [Lower Arm Length] | -0.23 | 1.83 | -3.82 | 3.36 | | 0.90 |
| Group [control]*Body Part [Neck Girth] | -0.32 | 0.99 | -2.26 | 1.63 | | 0.75 |
| Group [control]*Body Part [Neck Length] | -1.94 | 1.54 | -4.95 | 1.07 | | 0.21 |
| Group [control]*Body Part [Shoulder Width] | 1.21 | 1.96 | -2.63 | 5.05 | | 0.54 |
| Group [control]*Body Part [Abdomen Protrusion] | 5.05 | 2.60 | -0.05 | 10.14 | | 0.05 |
| Group [control]*Body Part [Thigh Girth] | 3.38 | 2.09 | -0.73 | 7.48 | | 0.11 |
| Group [control]*Body Part [Thigh Length] | 0.75 | 2.85 | -4.84 | 6.34 | | 0.79 |
| Group [control]*Body Part [Torso Height] | 3.87 | 3.82 | -3.62 | 11.37 | | 0.31 |
| Group [control]*Body Part [Upper Arm Girth] | 0.92 | 1.16 | -1.35 | 3.20 | | 0.43 |
| Group [control]*Body Part [Upper Arm Length] | 0.45 | 1.19 | -1.89 | 2.79 | | 0.71 |
| Group [control]*Body Part [Waist Size] | 2.44 | 2.62 | -2.71 | 7.58 | | 0.35 |
| Group [control]*Body Part [Wrist Girth] | - | - | - | - | | - |
| Height | -0.51 | 0.38 | -1.26 | 0.23 | | 0.18 |
| Weight | 0.59 | 0.48 | -0.36 | 1.54 | | 0.22 |
| BMI | -1.83 | 1.28 | -4.34 | 0.67 | | 0.15 |

*Notes:* Wrist girth set to zero since parameter is redundant. Height, weight, and BMI were covariates in the model.

Table S8: Women only parameter estimates for body dissatisfaction generalized estimating equation (GEE) with body size estimation (BSE) in the model

| *Predictors* | Estimates | Standard Error | 95% Confidence Interval | | | *p*-value |
| --- | --- | --- | --- | --- | --- | --- |
|  |  |  | Lower Bound | Upper Bound | |  |
| (Intercept) | -9.68 | 64.54 | -136.18 | | 116.82 | -9.68 |
| Group [control] | 0.83 | 1.14 | -1.40 | | 3.06 | 0.83 |
| BSE | -0.56 | 0.22 | -0.99 | | -0.13 | **0.01** |
| Group [control]*BSE | 0.10 | 0.28 | -0.45 | | 0.66 | 0.72 |
| Body Part [Ankle Girth] | 1.29 | 0.66 | -0.01 | | 2.59 | 0.05 |
| Body Part [Bust Girth] | 1.45 | 1.54 | -1.57 | | 4.47 | 0.35 |
| Body Part [Calf Girth] | -0.49 | 1.98 | -4.37 | | 3.40 | 0.81 |
| Body Part [Calf Length] | 8.14 | 1.80 | 4.60 | | 11.67 | **< 0.001** |
| Body Part [Chest Girth] | -8.22 | 2.47 | -13.07 | | -3.37 | **0.001** |
| Body Part [Foot Length] | 3.74 | 1.05 | 1.69 | | 5.79 | **< 0.001** |
| Body Part [Foot Width] | 1.81 | 0.89 | 0.06 | | 3.56 | **0.04** |
| Body Part [Hand Length] | 2.89 | 0.79 | 1.33 | | 4.45 | **< 0.001** |
| Body Part [Hand Width] | 1.93 | 0.77 | 0.43 | | 3.44 | **0.01** |
| Body Part [Hip Size] | -5.74 | 3.02 | -11.66 | | 0.19 | 0.06 |
| Body Part [Lower Arm Girth] | -2.21 | 0.52 | -3.23 | | -1.19 | **< 0.001** |
| Body Part [Lower Arm Length] | 1.44 | 1.25 | -1.01 | | 3.89 | 0.25 |
| Body Part [Neck Girth] | 0.11 | 0.74 | -1.35 | | 1.57 | 0.88 |
| Body Part [Neck Length] | 4.45 | 1.03 | 2.42 | | 6.47 | **< 0.001** |
| Body Part [Shoulder Width] | -3.70 | 1.60 | -6.84 | | -0.55 | **0.02** |
| Body Part [Abdomen Protrusion] | -1.96 | 1.94 | -5.76 | | 1.85 | 0.31 |
| Body Part [Thigh Girth] | -3.97 | 1.12 | -6.17 | | -1.78 | **< 0.001** |
| Body Part [Thigh Length] | 6.73 | 2.82 | 1.22 | | 12.25 | **0.02** |
| Body Part [Torso Height] | 6.70 | 2.28 | 2.23 | | 11.18 | **0.003** |
| Body Part [Upper Arm Girth] | -2.18 | 1.21 | -4.55 | | 0.19 | 0.07 |
| Body Part [Upper Arm Length] | 3.17 | 1.04 | 1.12 | | 5.21 | **0.002** |
| Body Part [Waist Size] | -9.51 | 2.13 | -13.70 | | -5.33 | **< 0.001** |
| Body Part [Wrist Girth] | - | - | - | | - | - |
| Group [control]*Body Part [Ankle Girth] | -1.68 | 1.22 | -4.07 | | 0.71 | 0.17 |
| Group [control]*Body Part [Bust Girth] | -0.70 | 2.22 | -5.06 | | 3.65 | 0.75 |
| Group [control]*Body Part [Calf Girth] | 1.09 | 2.74 | -4.28 | | 6.46 | 0.69 |
| Group [control]*Body Part [Calf Length] | -2.40 | 2.67 | -7.63 | | 2.82 | 0.37 |
| Group [control]*Body Part [Chest Girth] | 8.57 | 3.27 | 2.15 | | 14.98 | **0.01** |
| Group [control]*Body Part [Foot Length] | -1.53 | 1.40 | -4.28 | | 1.22 | 0.27 |
| Group [control]*Body Part [Foot Width] | -0.41 | 1.17 | -2.70 | | 1.89 | 0.73 |
| Group [control]*Body Part [Hand Length] | -0.96 | 1.10 | -3.12 | | 1.20 | 0.38 |
| Group [control]*Body Part [Hand Width] | -0.74 | 1.15 | -3.00 | | 1.51 | 0.52 |
| Group [control]*Body Part [Hip Size] | 6.28 | 3.82 | -1.20 | | 13.76 | 0.10 |
| Group [control]*Body Part [Lower Arm Girth] | 3.65 | 1.71 | 0.30 | | 7.00 | **0.03** |
| Group [control]*Body Part [Lower Arm Length] | -1.36 | 2.49 | -6.23 | | 3.52 | 0.59 |
| Group [control]*Body Part [Neck Girth] | 0.67 | 1.07 | -1.42 | | 2.76 | 0.53 |
| Group [control]*Body Part [Neck Length] | -2.39 | 1.60 | -5.53 | | 0.75 | 0.14 |
| Group [control]*Body Part [Shoulder Width] | 1.30 | 2.51 | -3.63 | | 6.23 | 0.61 |
| Group [control]*Body Part [Abdomen Protrusion] | 8.02 | 4.33 | -0.47 | | 16.51 | 0.06 |
| Group [control]*Body Part [Thigh Girth] | 2.85 | 1.86 | -0.78 | | 6.49 | 0.12 |
| Group [control]*Body Part [Thigh Length] | 2.10 | 3.80 | -5.34 | | 9.54 | 0.58 |
| Group [control]*Body Part [Torso Height] | 1.11 | 3.13 | -5.03 | | 7.25 | 0.72 |
| Group [control]*Body Part [Upper Arm Girth] | 2.13 | 1.69 | -1.19 | | 5.44 | 0.21 |
| Group [control]*Body Part [Upper Arm Length] | -0.47 | 1.55 | -3.50 | | 2.57 | 0.76 |
| Group [control]*Body Part [Waist Size] | 1.18 | 4.20 | -7.06 | | 9.41 | 0.78 |
| Group [control]*Body Part [Wrist Girth] | - | - | - | | - | - |
| Group [control]*Body Part [Ankle Girth]*BSE | 0.01 | 0.23 | -0.44 | | 0.46 | 0.97 |
| Group [control]*Body Part [Bust Girth]*BSE | 0.60 | 0.33 | -0.04 | | 1.24 | 0.07 |
| Group [control]*Body Part [Calf Girth]*BSE | 0.10 | 0.21 | -0.32 | | 0.52 | 0.65 |
| Group [control]*Body Part [Calf Length]*BSE | -0.18 | 0.24 | -0.65 | | 0.28 | 0.44 |
| Group [control]*Body Part [Chest Girth]*BSE | 0.34 | 0.16 | 0.03 | | 0.64 | **0.03** |
| Group [control]*Body Part [Foot Length]*BSE | 0.20 | 0.27 | -0.33 | | 0.73 | 0.47 |
| Group [control]*Body Part [Foot Width]*BSE | -0.08 | 0.23 | -0.53 | | 0.37 | 0.74 |
| Group [control]*Body Part [Hand Length]*BSE | -0.10 | 0.27 | -0.64 | | 0.43 | 0.71 |
| Group [control]*Body Part [Hand Width]*BSE | -0.44 | 0.36 | -1.16 | | 0.27 | 0.22 |
| Group [control]*Body Part [Hip Size]*BSE | 0.22 | 0.21 | -0.20 | | 0.63 | 0.31 |
| Group [control]*Body Part [Lower Arm Girth]*BSE | 0.67 | 0.25 | 0.19 | | 1.15 | **0.01** |
| Group [control]*Body Part [Lower Arm Length]*BSE | -0.11 | 0.31 | -0.71 | | 0.48 | 0.71 |
| Group [control]*Body Part [Neck Girth]*BSE | 0.59 | 0.19 | 0.22 | | 0.97 | **0.002** |
| Group [control]*Body Part [Neck Length]*BSE | 0.23 | 0.23 | -0.23 | | 0.68 | 0.33 |
| Group [control]*Body Part [Shoulder Width]*BSE | -0.20 | 0.26 | -0.71 | | 0.32 | 0.45 |
| Group [control]*Body Part [Abdomen Protrusion]*BSE | -0.19 | 0.39 | -0.96 | | 0.58 | 0.63 |
| Group [control]*Body Part [Thigh Girth]*BSE | -0.07 | 0.25 | -0.56 | | 0.42 | 0.79 |
| Group [control]*Body Part [Thigh Length]*BSE | 0.01 | 0.31 | -0.60 | | 0.61 | 0.98 |
| Group [control]*Body Part [Torso Height]*BSE | -0.06 | 0.26 | -0.57 | | 0.45 | 0.81 |
| Group [control]*Body Part [Upper Arm Girth]*BSE | 0.18 | 0.17 | -0.16 | | 0.51 | 0.31 |
| Group [control]*Body Part [Upper Arm Length]*BSE | 0.41 | 0.17 | 0.07 | | 0.75 | **0.02** |
| Group [control]*Body Part [Waist Size]*BSE | 0.20 | 0.22 | -0.23 | | 0.64 | 0.37 |
| Group [control]*Body Part [Wrist Girth]*BSE | - | - | - | | - | - |
| Height (m) | 0.09 | 0.40 | -0.69 | | 0.88 | 0.82 |
| Weight (kg) | -0.11 | 0.51 | -1.12 | | 0.89 | 0.83 |
| BMI (kg/m^2^) | -0.06 | 1.34 | -2.68 | | 2.57 | 0.97 |

*Notes:* Wrist girth set to zero since parameter is redundant. Height, weight, and BMI were covariates in the model.

Table S9: Women only GEE regression coefficients for BSE accuracy and body dissatisfaction predicting BDD symptom severity in the BDD group using the BDD-YBOCS and BABS

|  | **BDD-YBOCS** | | | | **BABS** | | | |
| --- | --- | --- | --- | --- | --- | --- | --- | --- |
| **Variable** | β | SE | 95% CI | *p* | β | SE | 95% CI | *p* |
| BSE Accuracy | -.001 | .03 | -.07, .07 | .97 | .05 | .02 | .004, .10 | .03 |
| Body Dissatisfaction | -.09 | .05 | -.18, .01 | .07 | -.02 | .04 | -.11, .06 | .61 |

Table S10: Women only GEE regression coefficients for BSE accuracy and body dissatisfaction predicting BDD symptom severity using the Body Image States Scale (BISS) across the full group

|  | **BISS** | | | |
| --- | --- | --- | --- | --- |
| **Variable** | β | SE | 95% CI | *p* |
| *Model 1* |  |  |  |  |
| BSE Accuracy | -.01 | .01 | -.03, .01 | .40 |
| Group | 2.65 | .30 | 2.06, 3.24 | **< .001** |
| Group*BSE Accuracy | .02 | .01 | -.01, .05 | .11 |
| *Model 2* |  |  |  |  |
| Body Dissatisfaction | .01 | .01 | -.02, .03 | .50 |
| Group | 2.72 | .39 | 1.95, 3.49 | **< .001** |
| Group*Body Dissatisfaction | -.02 | .02 | -.07, .02 | .36 |
